# Simple cumulative weighting of routine surveillance data identifies epidemic wave origins more accurately than a large language model: evidence from eight COVID-19 waves in Japan

**DOI:** 10.64898/2026.06.02.26354691

**Authors:** Shin-ichi Nakagawa, Akira Yamamoto

**Affiliations:** Research Institute of Info-Communication Medicine (RinCOM), Koganei, Tokyo, Japan; Faculty of Health Data Science, Juntendo University, Tokyo, Japan

**Keywords:** COVID-19, SARS-CoV-2, Epidemics, Disease Outbreaks, Population Surveillance, Forecasting, Spatial Analysis, Japan

## Abstract

Identifying the origin of an emerging epidemic wave within days of onset could enable targeted response before national spread, yet current methods rely on genomic sequencing that lags clinical detection by 2–4 weeks. We analysed daily COVID-19 cases from Japan’s 47 prefectures across eight waves (2020–2023), aggregated into 11 regional blocks. Wave onset was defined by the first difference of the K-value (K′). Six surveillance indicators were evaluated with and without cumulative historical weighting (λ = 0.75) and benchmarked against a large language model (Claude Haiku), scored by F1 against genomically confirmed origins. At 14 days after onset, cumulative weighting of peak and cumulative incidence (B1+prior, B3+prior) reached mean F1 = 0.622, exceeding the model (0.524); the gap was largest in Wave 7 (1.000 vs 0.333). Simple cumulative weighting of routine surveillance data identified wave origins more accurately than a language model, without proprietary tools or sequencing.

## Introduction

When a new epidemic wave begins, public health authorities face an urgent operational question: where did it start? Identifying the geographic origin within days of onset could allow targeted deployment of contact tracing, enhanced surveillance, and travel advisories before national spread renders region-specific intervention impractical (1,2). Definitive identification of wave origins currently relies on genomic surveillance, which typically lags clinical case detection by two to four weeks (3). Because origin-specific measures lose operational value once a wave has spread nationally, routine surveillance data able to identify the origin region within one to two weeks of onset—before genomic confirmation—would be of practical value.

Epidemic forecasting models based on fixed compartmental parameters, notably the SEIR framework, proved repeatedly inadequate during the COVID-19 pandemic, as they treat populations as homogeneous and assume stable transmission parameters—assumptions that break down when behavioural responses and variant-specific dynamics drive successive waves (4). This limitation motivated a shift toward data-driven approaches incorporating non-traditional behavioural signals. In Japan, deep learning models integrating mobile-phone footfall and social media activity as proxies for population behaviour projected epidemic wave peaks weeks to months in advance, substantially outperforming conventional approaches (5,6). A natural further question is whether large language models (LLMs)—which integrate qualitative epidemiological knowledge, variant characteristics, and historical outbreak patterns from their training data— could extend this capability to identifying the geographic origin of an emerging wave in real time (13,14). This hypothesis has not been directly tested at the subnational level.

A further methodological gap concerns the definition of wave onset. Conventional spatial outbreak-detection approaches—cross-correlation analysis, Granger causality testing, and growth-rate metrics—treat each wave independently and depend critically on when the analysis window opens (7,8). If onset is defined by administrative boundaries rather than epidemiological data, early-phase observations may include residual transmission from the preceding wave, contaminating the origin signal with noise from a geographically distinct pattern. Despite growing interest in systematic epidemic wave characterisation (11), the sensitivity of origin identification to onset definition has not been evaluated, nor have cumulative and non-cumulative analytical approaches been compared within a consistent prospective framework across multiple successive waves.

We address these gaps using daily COVID-19 confirmed case data from Japan’s 47 prefectures across eight epidemic waves (2020–2023), aggregated into 11 regional blocks. We propose a data-driven wave onset definition based on the first difference of the K-value (K′) (9), which detects sustained acceleration of epidemic growth velocity and separates residual transmission from the preceding wave from the genuine early-phase signal of the new wave. We systematically evaluate six candidate statistical indicators under this framework, with and without cumulative historical weighting, and benchmark their performance against a large language model provided with identical historical context. The primary outcome is F1 score against genomically confirmed regional origins, evaluated at 7, 14, 21, and 28 days after wave onset across Waves 2–8.

## Methods

### Data sources

Daily prefecture-level COVID-19 confirmed case counts were obtained from the Ministry of Health, Labour and Welfare (MHLW) open data portal, spanning January 2020 through May 2023 (10). Population-adjusted rates (cases per 100,000 population) were calculated using 2020 national census denominators. All data were smoothed using a 7-day centred moving average before analysis.

### Wave onset definition

Wave onset was defined using a two-step, data-driven procedure based on the K-value, a dimensionless indicator of epidemic growth velocity (9). The K-value on day d is K(d) = [N(d) − N(d−7)] / N(d), where N(d) is cumulative confirmed cases on day d, representing the proportion of cumulative cases attributable to the preceding seven days. To reduce day-to-day noise, K(d) was first smoothed with a 7-day trailing moving average, K_0_(d) = mean[K(d−6),…,K(d)]. The first difference K′(d) = K_0_(d) − K_0_(d−1) captures the rate of change of smoothed growth velocity, with positive values indicating sustained acceleration of spread.

For each of the 11 regional blocks, the monitoring window for wave n opened on the administrative start date of wave n. Block-level onset was defined as the first day within this window on which K′ had remained positive for 14 consecutive days; the wave-level onset date was the earliest block-level onset across all 11 blocks. A sensitivity analysis comparing thresholds of 7, 14, and 21 consecutive days confirmed that 14 days optimised the balance between detection stability and predictive performance (Supplementary Table S3). To further validate the criterion, we compared it against two alternatives: effective reproduction number Rt ≥ 1 sustained for 4 consecutive days (serial interval = 5 days), and an absolute threshold of 0.3 cases/100,000 (7-day trailing moving average). K′ with n = 14 outperformed both in downstream origin-identification F1: the 0.3/100K threshold failed for Waves 7 and 8, where inter-wave troughs remained above threshold throughout, and the Rt-based approach yielded lower mean F1 across most transitions (Supplementary Figure S4).

A fully prospective definition—determining both onset and termination from real-time data alone—was explored but proved infeasible. Wave termination in particular is difficult to detect prospectively, as transient decreases in K′ frequently occur during a wave’s declining phase without representing true termination, and no data-driven termination criterion proved stable across all eight waves. The hybrid approach adopted here retains the epidemiological validity of the K′-based onset criterion while avoiding this instability.

### Regional block definition

Japan’s 47 prefectures were aggregated into 11 regional blocks based on geographic proximity and established public health administrative boundaries: Hokkaido, Tohoku, Kanto, Koshinetsu, Hokuriku, Tokai, Kinki, Chugoku, Shikoku, Kyushu, and Okinawa. Regional incidence rates were calculated as unweighted means of constituent prefecture rates derived from the 7-day centred moving average of daily confirmed cases per 100,000 population.

### Candidate indicators

We evaluated six candidate indicators (B0–B5) derivable solely from routine surveillance data, spanning four conceptual categories: onset timing (B0, earliest block-level onset day); magnitude (B1, peak incidence rate; B3, cumulative incidence rate); growth dynamics (B2, OLS growth rate); and spatial relationships (B4, cross-correlation lead score; B5, Granger causality score). All indicators were computed from block-level data from wave onset through day N (N = 7, 14, 21, 28) and min–max normalised to [0, 1] across the 11 blocks before ranking. Complete mathematical definitions are provided in the Supplementary Appendix.

### Cumulative historical weighting

For each target wave n, an additive cumulative score was computed as Score(r) = Score_baseline(r) + λ × P(r, n), where P(r, n) is the proportion of prior waves (1 through n−1) in which region r was a genomically confirmed origin and λ = 0.75, selected by sensitivity analysis across λ ∈ {0, 0.25, 0.50, 0.75, 1.00} (Supplementary Table S1). This formulation ensures strict temporal separation: wave n contributes no information to its own prediction. Cumulative weighting was applied to the three quantitative indicators (yielding B1+prior, B2+prior, B3+prior); B0, B4, and B5 showed no meaningful improvement in preliminary analyses and were retained in non-cumulative form.

### LLM benchmark

Claude Haiku (claude-haiku-4-5-20251001; Anthropic) was evaluated without fine-tuning under a cumulative historical context condition. For each wave–timepoint combination, the model received a structured prompt comprising the confirmed genomic origins of all prior waves (1 through n−1) by variant, and current-wave block-level data for the first N days—cumulative incidence rate, peak incidence rate, and days since onset, ranked in descending order of cumulative incidence. The model was instructed to identify the top-3 origin blocks in JSON format. Three independent runs were conducted per combination; mean F1 scores are reported. The complete prompt structure is provided in the Supplementary Appendix.

### Performance evaluation

Performance was assessed by comparing the top-3 predicted origin blocks against genomically confirmed regional origins for each wave (Table 2). For each prediction we calculated precision (proportion of predicted blocks that were confirmed origins), recall (proportion of confirmed origins that were predicted), and F1 (harmonic mean of precision and recall). F1 ranges from 0 to 1 and penalises false positives and false negatives equally, making it appropriate where both over- and under-prediction carry operational cost. All statistical methods predicted exactly three origin blocks. Mean F1 scores were computed as unweighted averages across Waves 2–8 (seven waves). All analyses were performed in Python 3.12.

## Results

### Wave onset dates under the K′-based definition

Application of the K′-based onset definition yielded wave-level onset dates ranging from 1 June 2020 (Wave 2, Kanto) to 13 October 2022 (Wave 8, Hokkaido). The inter-block spread of onset dates varied markedly across waves, reflecting differences in epidemic structure (Figure 1A). Wave 5 (Delta) showed the most spatially concentrated onset: all 11 blocks reached the K′ criterion on the same day (1 August 2021), providing no spatial differentiation and explaining the near-zero performance of growth-dynamic indicators (B2, B4) for this wave. Wave 2 (Ancestral) showed the greatest geographic spread, with Kanto reaching onset first on 1 June 2020, followed by Hokkaido 109 days later—consistent with the limited inter-regional mobility of the early pandemic period. Waves 3 and 4 showed near-simultaneous onset across most blocks (8 of 11 within 3 days for Wave 3; 9 of 11 for Wave 4), reflecting rapid national spread. Wave 6 (Omicron BA.1) was notable for a structural mismatch between the earliest-detected block (Koshinetsu, 3 December 2021) and the genomically confirmed origins (Okinawa, Chugoku), which reached the K′ threshold 13 and 12 days later, respectively (Figure 1B). This mismatch contributed to the uniformly low F1 scores observed across all methods for this wave. Wave 7 (Omicron BA.2/5) similarly showed onset first in Kyushu (13 June 2022), 12 days before the confirmed origin blocks (Kanto, Okinawa, Kinki), yet cumulative historical weighting correctly recovered the confirmed origins by elevating these historically frequent blocks above the early-onset signal.

**Figure 1.**
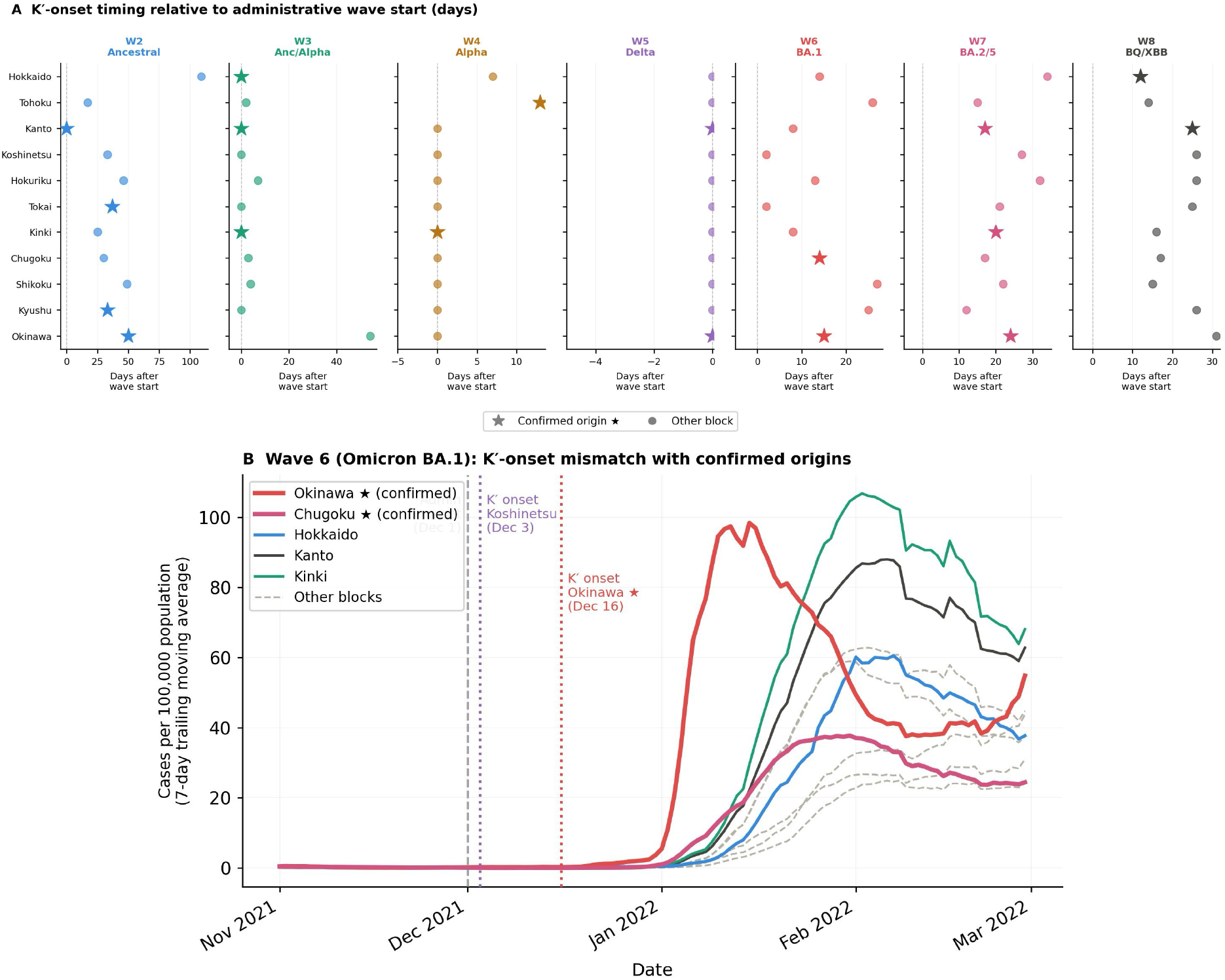
K′-based wave onset across 11 regional blocks. (A) K′-onset timing relative to administrative wave start for seven epidemic waves (W2–W8). Stars (⋆) mark genomically confirmed origin blocks; circles mark other blocks. Wave 5 (Delta) shows simultaneous onset across all 11 blocks (all points at day 0); Wave 2 shows the widest inter-block spread (109 days). (B) Wave 6 (Omicron BA.1) incidence time series illustrating the structural mismatch between K′-detected onset and confirmed origins. Vertical lines mark the administrative onset (1 December 2021), the earliest K′ onset (Koshinetsu, 3 December 2021), and the confirmed-origin K′ onset (Okinawa, 16 December 2021).

### Performance of non-cumulative statistical indicators

At 14 days after wave onset, non-cumulative statistical indicators achieved mean F1 scores ranging from 0.250 (B5, Granger causality) to 0.510 (B1, peak incidence rate; B3, cumulative incidence rate) (Table 1, Figure 2). B1 and B3 were the strongest non-cumulative indicators, each at F1 = 0.510, followed by B0 (earliest onset day, F1 = 0.412). B2 (OLS growth rate, F1 = 0.244), B4 (cross-correlation, F1 = 0.307), and B5 (F1 = 0.250) were consistently lower.

**Table 1.**
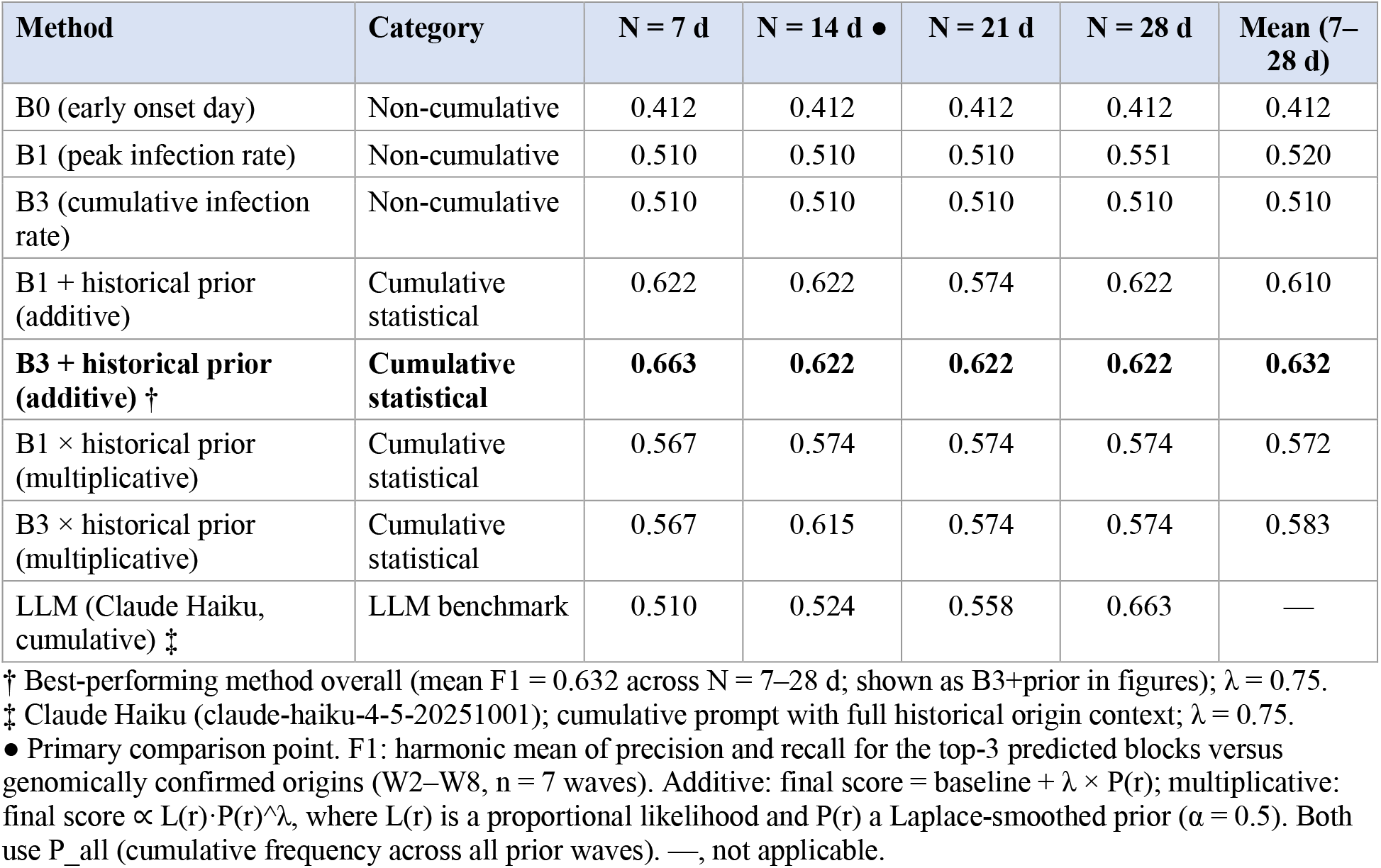
Performance of origin-identification methods across observation windows.

| Method | Category | N = 7 d | N = 14 d • | N = 21 d | N = 28 d | Mean (7–28 d) |
| --- | --- | --- | --- | --- | --- | --- |
| B0 (early onset day) | Non-cumulative | 0.412 | 0.412 | 0.412 | 0.412 | 0.412 |
| B1 (peak infection rate) | Non-cumulative | 0.510 | 0.510 | 0.510 | 0.551 | 0.520 |
| B3 (cumulative infection rate) | Non-cumulative | 0.510 | 0.510 | 0.510 | 0.510 | 0.510 |
| B1 + historical prior (additive) | Cumulative statistical | 0.622 | 0.622 | 0.574 | 0.622 | 0.610 |
| **B3 + historical prior (additive) †** | **Cumulative statistical** | **0.663** | **0.622** | **0.622** | **0.622** | **0.632** |
| B1 × historical prior (multiplicative) | Cumulative statistical | 0.567 | 0.574 | 0.574 | 0.574 | 0.572 |
| B3 × historical prior (multiplicative) | Cumulative statistical | 0.567 | 0.615 | 0.574 | 0.574 | 0.583 |
| LLM (Claude Haiku, cumulative) ‡ | LLM benchmark | 0.510 | 0.524 | 0.558 | 0.663 | — |

† Best-performing method overall (mean F1 = 0.632 across N = 7–28 d; shown as B3+prior in figures); $\lambda = 0.75$ .
‡ Claude Haiku (claude-haiku-4-5-20251001); cumulative prompt with full historical origin context; $\lambda = 0.75$ .
• Primary comparison point. F1: harmonic mean of precision and recall for the top-3 predicted blocks versus genomically confirmed origins (W2–W8, n = 7 waves). Additive: final score = baseline + $\lambda \times P(r)$ ; multiplicative: final score $\propto L(r) \cdot P(r)^\lambda$ , where $L(r)$ is a proportional likelihood and $P(r)$ a Laplace-smoothed prior ( $\alpha = 0.5$ ). Both use $P_{\text{all}}$ (cumulative frequency across all prior waves). —, not applicable.

**Figure 2.**
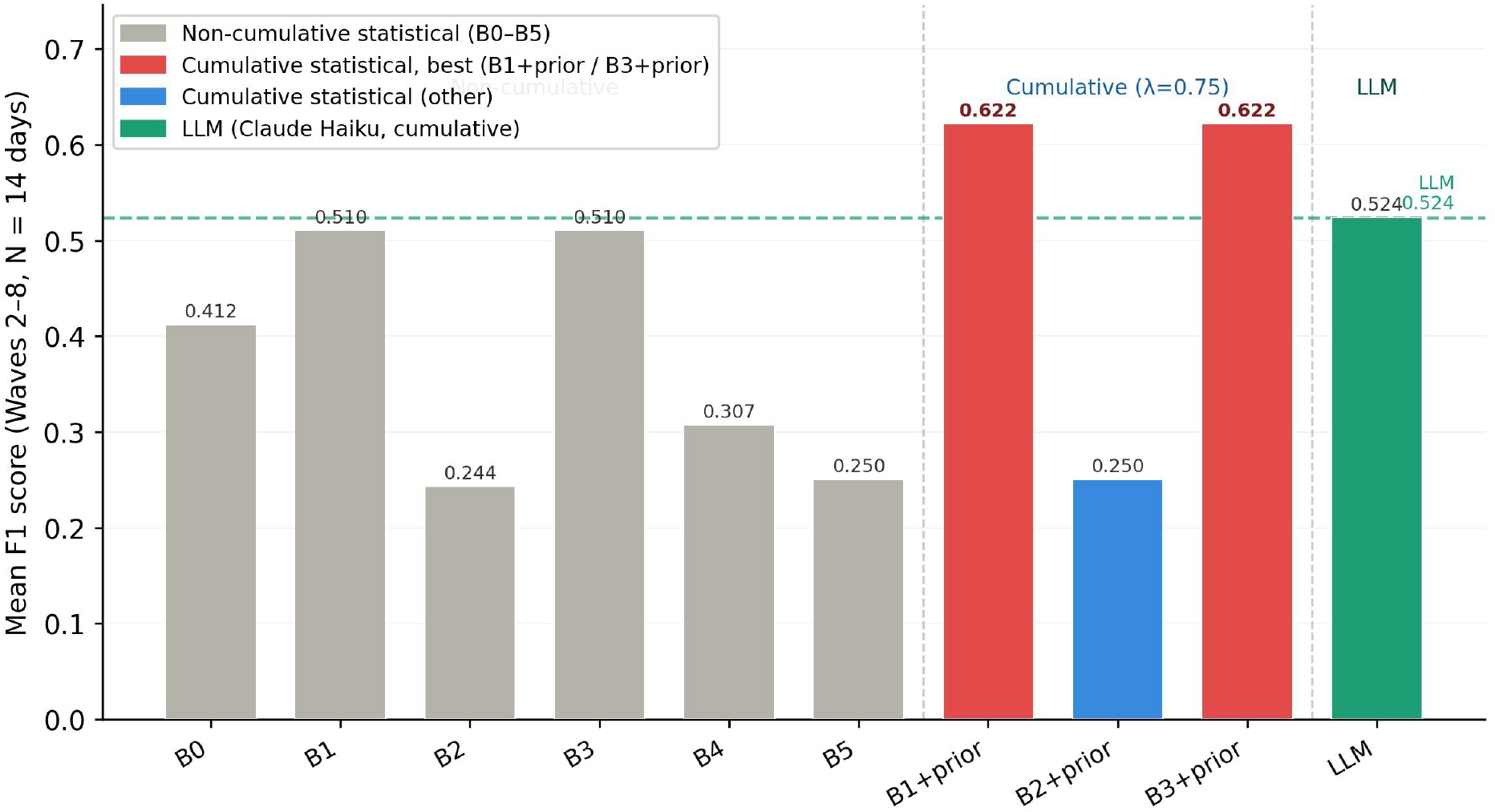
Mean F1 at N = 14 days after wave onset for all methods (Waves 2–8). Bars show six non-cumulative indicators (B0–B5), three additive cumulative indicators (B1+prior, B2+prior, B3+prior), and the cumulative LLM. The dashed line marks the cumulative LLM (F1 = 0.524). B1+prior and B3+prior (F1 = 0.622) exceed all other methods, including the LLM.

Wave-specific analysis showed that the magnitude indicators reached F1 = 0.800 in Wave 5 (Delta) and 0.571 in Wave 2 (Ancestral), but were limited to F1 = 0.333 in Wave 7 (Omicron BA.2/5) and 0.400 in Waves 4, 6, and 8 (Table 2). Wave 5’s high performance reflects the clear magnitude gradient between confirmed origin blocks (Okinawa, Kanto) and other regions at onset, whereas Wave 7’s low non-cumulative performance reflects the mismatch between onset-period incidence and the historically established origin blocks. Performance was stable across time points: mean F1 at N = 7, 14, and 21 days differed by ≤ 0.06 for B1 and B3, indicating that origin signals are detectable within the first week and do not substantially improve with additional data under the non-cumulative framework.

**Table 2.** Wave-specific F1 scores at N = 14 days (primary comparison point).

| Wave | Dominant variant | B3 | B1 + prior (additive) | B3 + prior (additive) † | LLM ‡ |
| --- | --- | --- | --- | --- | --- |
| W2 | Ancestral | 0.571 | 0.286 | 0.286 | 0.667 |
| W3 | Anc/Alpha | 0.667 | 0.667 | 0.667 | 0.667 |
| W4 | Alpha | 0.400 | 0.400 | 0.400 | 0.400 |
| W5 | Delta | 0.800 | 0.800 | 0.800 | 0.800 |
| W6 ‖ | Omicron BA.1 | 0.400 | 0.400 | 0.400 | 0.400 |
| W7 ** | Omicron BA.2/5 | 0.333 | **1.000** | **1.000** | 0.333 |
| W8 | Omicron BQ/XBB | 0.400 | 0.800 | 0.800 | 0.400 |
| **Mean** | — | **0.510** | **0.622** | **0.622** | **0.524** |

† B3+prior in figures; $\lambda = 0.75$ . ‡ LLM (Claude Haiku, claude-haiku-4-5-20251001); cumulative prompt with historical origin context; mean F1 = 0.524 across W2–W8.
‖ Wave 6 (Omicron BA.1): all methods fail to exceed F1 = 0.400, owing to a structural mismatch between K'-detected onset and genomically confirmed origins (see Discussion).
\*\* Wave 7 (Omicron BA.2/5): cumulative statistical methods achieve perfect identification (F1 = 1.000) while the LLM does not (F1 = 0.333), illustrating the advantage of explicit historical-prior weighting when current-wave evidence is ambiguous.

### Effect of cumulative historical weighting

Addition of cumulative historical weighting (λ = 0.75) improved mean F1 for the magnitude indicators from 0.510 to 0.622 at N = 14 days (Table 1). B2+prior showed no meaningful improvement (0.244 → 0.250), consistent with the poor baseline performance of the growth-rate indicator.

The gain was concentrated in two waves. In Wave 7 (Omicron BA.2/5), weighting elevated Kanto, Okinawa, and Kinki—the three most frequently confirmed origin blocks across prior waves—above the early-onset signal from Kyushu and Tohoku, raising F1 from 0.333 to 1.000. In Wave 8 (Omicron BQ/XBB), it raised F1 from 0.400 to 0.800 by correctly elevating Hokkaido and Kanto. Weighting was neutral for Waves 3–6. In Wave 2, however, where only Wave 1 was available as historical context, the prior was misleading and reduced F1 from 0.571 to 0.286 (Table 2)—an early-wave limitation we revisit below.

The λ sensitivity analysis confirmed λ = 0.75 as optimal; leave-one-wave-out analysis found λ ≥ 0.75 optimal in 6 of 7 waves, the sole exception being Wave 7, where λ = 0.50 was marginally preferred on the remaining-six-wave criterion although held-out F1 was 1.000 at both values (Supplementary Tables S1, S2).

### LLM benchmark

The cumulative LLM (Claude Haiku) reached mean F1 = 0.524 at N = 14 days, below both B1+prior and B3+prior (F1 = 0.622) (Table 1, Figure 2). The difference was most pronounced in Wave 7, where the statistical methods achieved F1 = 1.000 and the LLM 0.333 (Table 2).

Despite receiving identical historical origin information, the LLM did not systematically elevate the historically frequent origin blocks (Kanto, Okinawa, Kinki) above the early-onset signal from Kyushu and Tohoku, which suggests that qualitative contextual reasoning is less reliable than explicit arithmetic weighting for this class of structured prediction task.

Conversely, the LLM outperformed the statistical methods in Wave 2 (F1 = 0.667 vs 0.286), where only one prior wave was available as context. In this data-sparse setting, the model’s broader epidemiological priors—knowledge of Kanto (the Tokyo metropolitan area) as Japan’s principal international gateway—appear to have supplied a signal the single-wave arithmetic prior could not. LLM performance also improved with accumulating data, rising from F1 = 0.510 at N = 7 to 0.663 at N = 28 and surpassing both statistical methods (0.622) by N = 28 (Figure 3). This trajectory suggests that LLMs may become more competitive as within-wave data accumulate, although the advantage of simple cumulative weighting is greatest at the early time points when rapid decisions are required.

**Figure 3.**
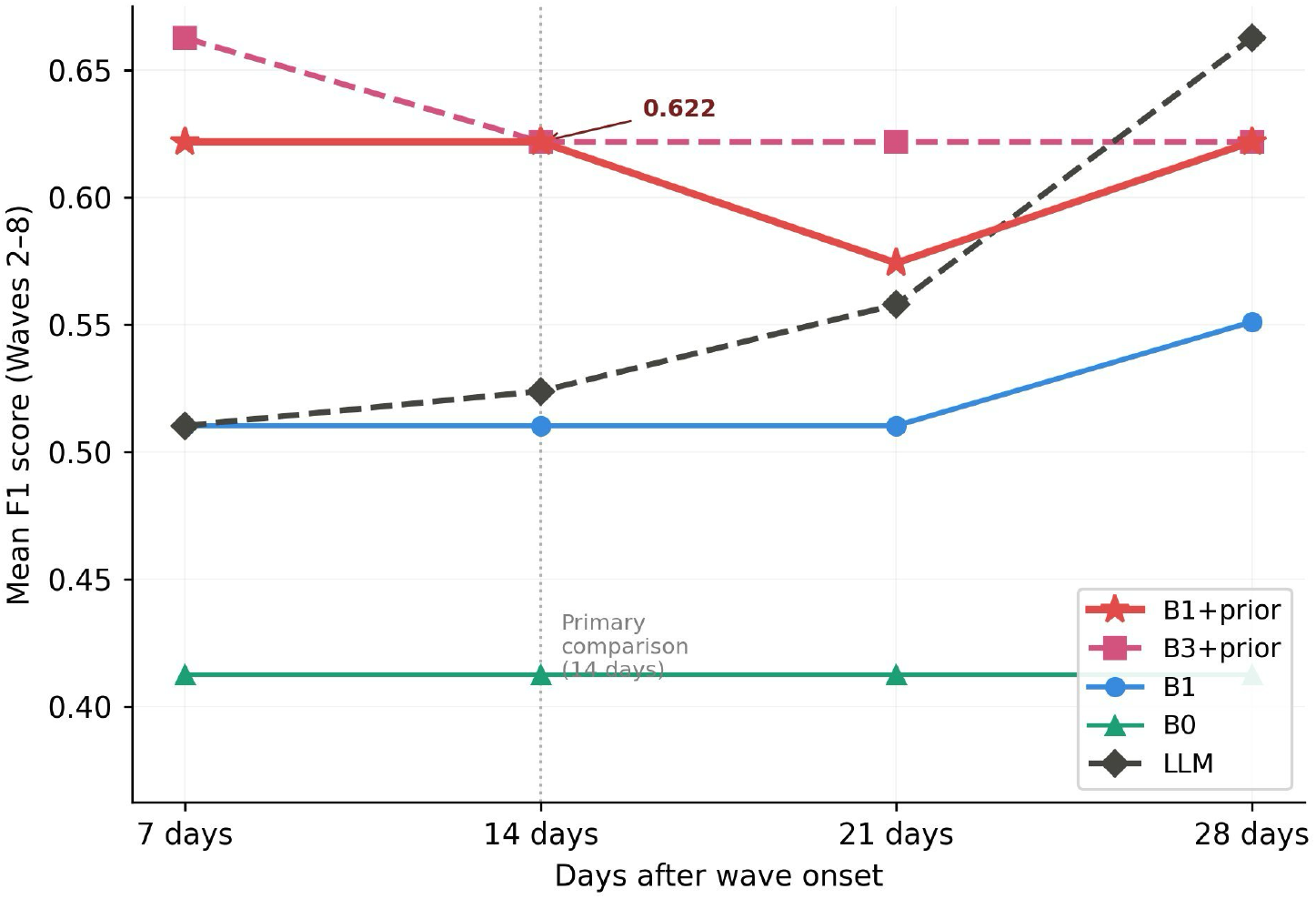
Mean F1 as a function of observation window (N = 7, 14, 21, 28 days) for key methods (Waves 2–8). At 14 days, B1+prior and B3+prior (F1 = 0.622) exceed the LLM (0.524). The ranking reverses by N = 28 days, where the LLM (F1 = 0.663) surpasses B3+prior (0.622), indicating that the statistical advantage is greatest at early time points when data are limited and decisions most time-sensitive.

## Discussion

### A data-driven onset definition as a prerequisite for origin identification

A central methodological observation of this study is that wave onset definition is itself a primary determinant of indicator performance. The K′-based definition—requiring 14 consecutive days of accelerating epidemic growth velocity—separates residual transmission from the preceding wave from the genuine early-phase signal of the emerging wave. This separation appears essential: when onset is defined administratively, early-phase data may include residual cases from the prior wave, obscuring the origin signal. The sensitivity analysis confirmed that 14 days optimised the balance between detection stability and predictive performance (Supplementary Table S3), with 21 days producing unacceptable detection failures in early waves when cumulative case counts were low.

The wave-specific onset patterns carry epidemiological meaning in their own right. Wave 5 (Delta) showed simultaneous onset across all 11 blocks on a single day, consistent with the rapid, spatially uniform acceleration of Delta and the near-complete depletion of susceptibles in high-contact populations. Wave 2, by contrast, showed a 109-day spread between the earliest (Kanto) and latest (Hokkaido) block onsets, reflecting the restricted inter-regional mobility of the early pandemic period. These differences suggest that no single onset definition can be assumed to generalise, and that monitoring the K′ signal in real time offers a wave-specific, data-driven alternative to fixed administrative boundaries. Monitoring K′ in real time additionally yields interpretable onset chronologies: the contrast between Wave 5’s single-day, all-block onset and Wave 2’s 109-day cascade is itself a descriptive output of operational interest, independent of the origin-identification task.

### Arithmetic cumulative weighting and LLM-based contextual reasoning

The central finding is that a simple arithmetic operation—adding a historically derived prior to a normalised surveillance score—identified wave origins more accurately than a state-of-the-art large language model at the operationally critical 14-day point (mean F1 = 0.622 vs 0.524). Both methods received identical historical information: the statistical methods translated it into an explicit numerical adjustment (λ × P(r, n)) applied uniformly across blocks, whereas the LLM processed it through natural-language reasoning and tended to anchor predictions on the early-onset incidence pattern rather than the historical prior. This pattern is consistent with a recency bias in structured quantitative tasks: when current data conflicted with the historical prior, the LLM appeared to weight the current signal more heavily than the arithmetic method did. The practical implication is that B1+prior and B3+prior require only routinely available incidence data, a list of historically confirmed origins, and a spreadsheet; they are auditable, deterministic, and require no proprietary model access—properties of operational value where explainability, reproducibility, and resource constraints matter. More broadly, these results do not argue against language models in public health but help delineate where they add value: for this narrowly structured task—mapping a fixed historical frequency onto a current ranking—an explicit arithmetic rule proved more reliable and transparent than free-form reasoning, whereas the model’s general knowledge was most useful precisely where structured historical data were too sparse to compute a stable prior.

To assess whether the additive form was merely a convenient approximation of a more principled probabilistic model, we reformulated cumulative weighting as a conditional probability, final score ∝ L(r)·P(r)^λ, where L(r) is a proportional likelihood from normalised surveillance indicators and P(r) is a Laplace-smoothed empirical prior over historical origin frequencies, re-optimising λ independently from 0.10 to 2.00. The additive form retained a marginal advantage (B3+prior, λ = 0.75, mean F1 = 0.632 versus the multiplicative B3 at optimal λ = 1.00, mean F1 = 0.583), and the cumulative prior (P_all) consistently outperformed a prior based on the immediately preceding wave alone (P_prev). These results suggest the additive formulation is not merely a convenient approximation but an empirically sufficient representation of the underlying conditional structure.

This interpretation is reinforced by the one wave in which the LLM prevailed. In Wave 2, with only a single prior wave available, the arithmetic prior was both sparse and, in this instance, misleading, reducing F1 from 0.571 to 0.286; the LLM, drawing on general epidemiological knowledge rather than a one-sample frequency, correctly favoured Kanto. The cumulative-weighting advantage therefore depends on a sufficient historical record and is expected to strengthen as more waves accrue—the regime in which the method is intended to operate.

### Wave 6 as a structural exception

Wave 6 (Omicron BA.1) was the most informative failure case. All methods—cumulative and non-cumulative alike—reached only F1 = 0.400. The uniform failure reflects a structural mismatch between the K′-detected onset pattern and the confirmed origins: Koshinetsu and Tokai showed the earliest accelerating K′ signal, while Okinawa and Chugoku—the confirmed origins—reached the onset threshold 13 and 12 days later, respectively (Figure 1B). This is consistent with the atypical introduction pathway of Omicron BA.1, which entered through international travel channels during a period of border restrictions, seeding regions with international connectivity while K′ detected earlier community-level acceleration elsewhere. A systematic divergence between the K′-based onset pattern and the cumulative historical prior may itself serve as an operational signal for atypical variant introductions warranting enhanced genomic surveillance. That the same cumulative prior recovered the correct origins in the immediately following Wave 8 (F1 = 0.400 → 0.800) indicates that the Wave 6 failure was specific to this introduction event rather than a persistent breakdown of the framework.

### Limitations

Several limitations should be noted. First, the evaluation covers eight waves in a single country with a specific surveillance infrastructure; external validation across different pathogens, surveillance systems, and geographic scales is essential before generalising. Second, although a multiplicative conditional-probability reformulation was evaluated and the additive form proved empirically sufficient (above), formal Bayesian model comparison remains warranted. Third, the LLM evaluated here represents one model at one point in time; more capable models or refined prompting strategies may yield different results. Fourth, the top-3 prediction rule, λ = 0.75, and the 14-day onset threshold were each selected through internal analyses rather than independent external validation. Fifth, block-level incidence was computed as unweighted means of constituent prefecture rates, which does not account for differences in prefecture population size within blocks.

## Conclusions

Simple cumulative weighting of routine surveillance data identified epidemic wave origins more accurately than a state-of-the-art large language model at the operationally relevant time point, and can be implemented without proprietary tools—an advantage for resource-constrained public health settings. The K′-based onset definition, and the divergence between early-onset patterns and historical priors, may together serve as operational signals for atypical variant introductions warranting enhanced genomic surveillance. Combined with our companion analysis of next-wave magnitude (12), this framework may help anticipate both where a future epidemic wave will begin and how large it will become, using only data available at wave onset.

## Data Availability

Prefecture-level COVID-19 case data are available from the Ministry of Health, Labour and Welfare of Japan open data portal (https://www.mhlw.go.jp/stf/covid-19/open-data.html). Analysis code is available from the corresponding author upon reasonable request.

## Acknowledgments

We thank the Ministry of Health, Labour and Welfare of Japan for making prefecture-level COVID-19 surveillance data publicly available.

## AI use disclosure

The authors used Claude (Anthropic, claude.ai), an LLM-based AI assistant, to support data analysis, manuscript drafting, figure generation, and coding throughout this study. All analytical results, interpretations, and conclusions were verified and approved by the authors. Claude Haiku (claude-haiku-4-5-20251001) was additionally evaluated as the benchmark large language model, as described in Methods.

## Biographical sketch

Dr. Nakagawa is a physician-researcher at the Research Institute of Info-Communication Medicine (RinCOM), Koganei, Tokyo, Japan. His research integrates routine surveillance data, statistical methods, and large language models for real-time epidemiological analysis.

## Supplementary Appendix

### Supplementary Methods: Complete Indicator Definitions (B0–B5)

All indicators were evaluated using data from wave onset through day N (N = 7, 14, 21, 28 days after K′-onset confirmation). Scores were min-max normalised to [0, 1] across all 11 blocks; top-3 blocks designated as predicted origins.

#### B0 – Earliest Onset Day

Block for which K′-onset (14 consecutive days of K′ > 0) is confirmed earliest after the administrative wave start.

#### B1 – Peak Infection Rate

Maximum 7-day trailing moving-average incidence during the first N days after K′-onset confirmation.

#### B2 – OLS Growth Rate

OLS slope of the N-day incidence series, normalised by mean incidence.

#### B3 – Cumulative Infection Rate

Sum of smoothed daily incidence over the first N days after K′-onset confirmation.

#### B4 – Cross-correlation Lead Score

Proportion of blocks for which r’s series precedes the other block based on lag maximising cross-correlation.

#### B5 – Granger Causality Score

Proportion of blocks for which r Granger-causes the other block at p < 0.05 (bivariate VAR, lag = 3).

### Supplementary Methods: Cumulative Historical Weighting

Additive formulation: Score_cumul(r) = Score_baseline(r) + λ × P(r, n), where P(r, n) = [1/(n−1)]Σ I(r ∈ G_k) is the proportion of prior waves in which block r was a confirmed origin, and λ = 0.75 (see Table S1).

A multiplicative formulation (Score_mul(r) ∝ L(r)·P(r)^λ, Laplace-smoothed prior α = 0.5) was also evaluated; the additive formulation retained a marginal advantage for primary indicators (see main text).

**Table S1.** Sensitivity analysis: effect of λ on mean F1 (B3 + prior, Waves 2–8)

| $\lambda$ | W3 | W5 | W7 | W8 | Mean (W2–8, N=7–28 d) |
| --- | --- | --- | --- | --- | --- |
| 0.00 (no weighting) | 0.667 | 0.800 | 0.333 | 0.400 | 0.510 |
| 0.25 | — | — | — | — | 0.539 |
| 0.50 | — | — | — | — | 0.580 |
| **0.75 (selected)** | **0.667** | **0.800** | **1.000** | **0.800** | **0.632** |
| 1.00 | — | — | — | — | 0.622 |
| 1.50 | — | — | — | — | 0.622 |

$\lambda$ : historical weighting parameter. Mean F1 across N = 7–28 d, Waves 2–8. Wave-by-wave values shown for $\lambda = 0.00$ and 0.75 at N = 14 only. Green: selected ( $\lambda = 0.75$ ).

**Table S2.**
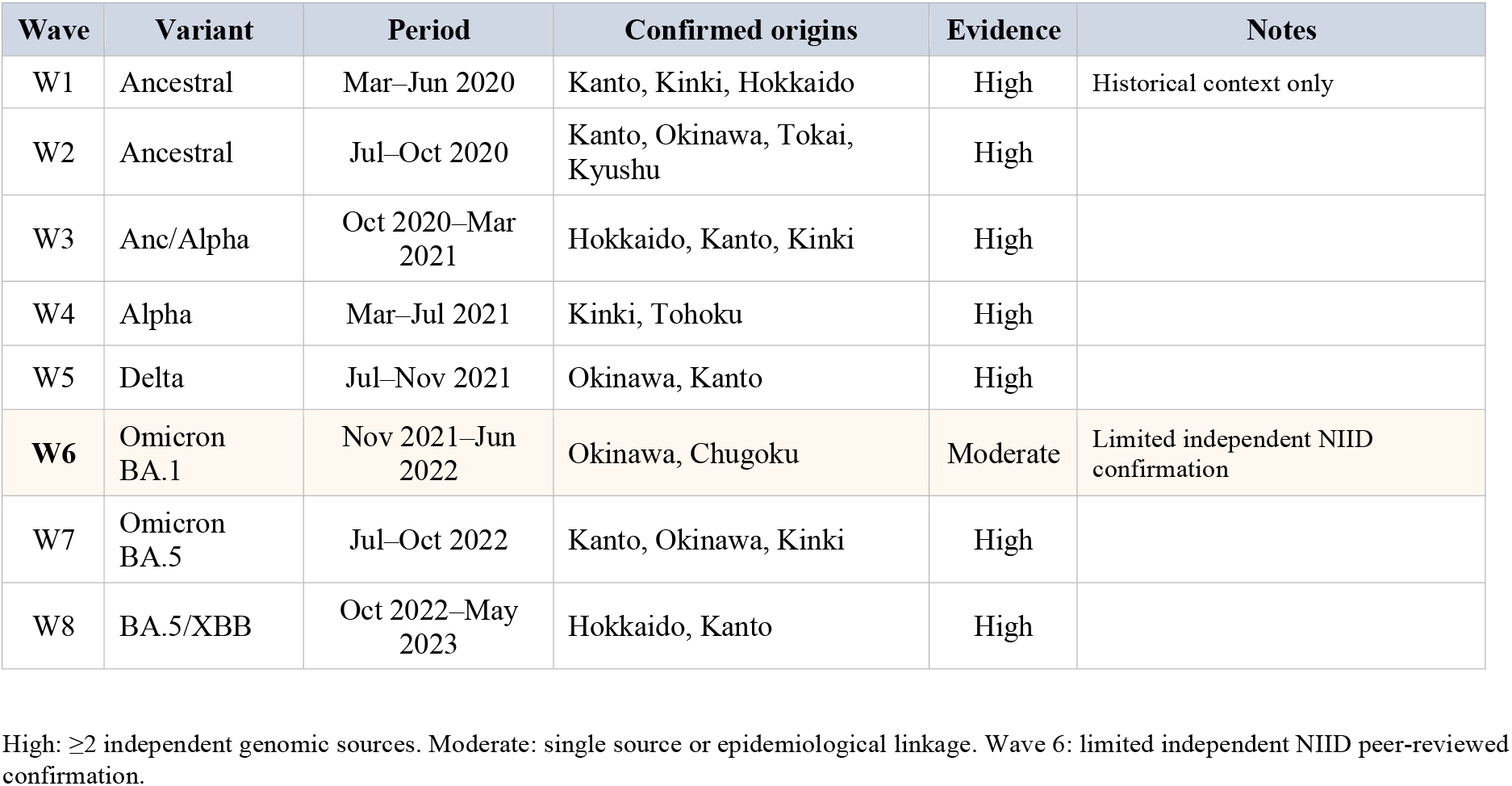
Genomically confirmed wave origins used as ground truth.

**Table S3.** Sensitivity analysis: effect of K′-onset threshold (n_onset) on mean F1 at N = 14 days.

| Method | n_onset = 7 d | n_onset = 14 d (selected) | n_onset = 21 d |
| --- | --- | --- | --- |
| B0 | **0.469** | 0.412 | 0.422 |
| B1 | **0.567** | 0.510 | 0.453 |
| B2 | 0.219 | **0.301** | 0.253 |
| B3 | **0.567** | 0.510 | 0.453 |
| B4 | 0.298 | 0.355 | **0.412** |
| B5 | **0.203** | 0.203 | 0.203 |
| **B1+prior** | **0.622** | 0.622 | 0.605 |
| **B2+prior** | **0.307** | 0.250 | 0.260 |
| **B3+prior** | **0.622** | 0.622 | 0.605 |
| Detection failures† | 0 / 0 | 2 / 0 | 9 / 4 |

Blue: selected (n\_onset = 14). Green: maximum F1 for that method. †Detection failures: blocks (of 11) for Wave 1 / Wave 2. n\_onset = 21 yielded 9/4 failures, rendering it unsuitable.

**Figure S4.**
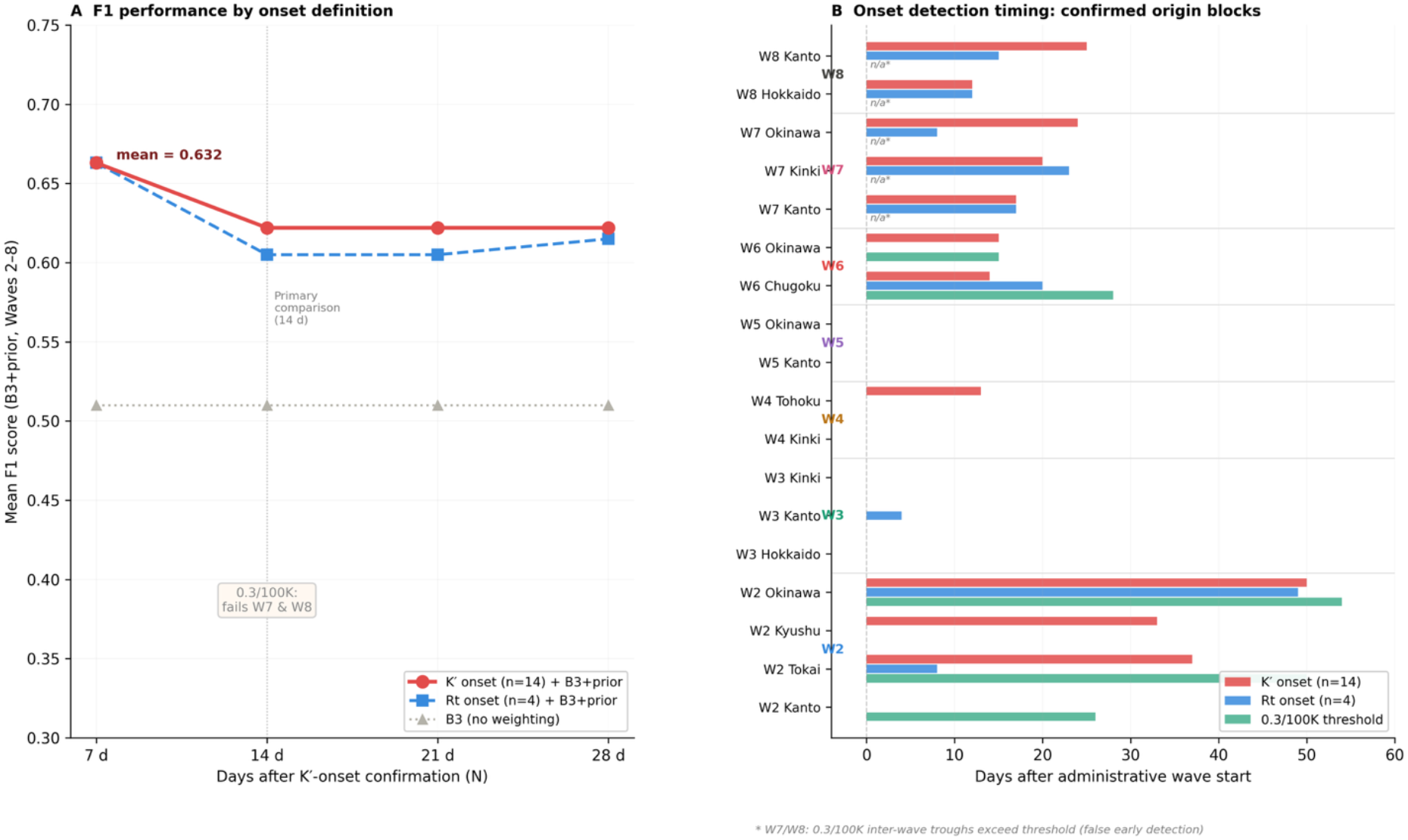
Comparison of three wave-onset definitions. (A) Mean F1 (B3+prior, Waves 2–8) under K′ onset (selected), Rt onset (Rt ≥ 1, 4 d), and B3 without weighting, vs observation window N. (B) Days from administrative wave start to onset detection for confirmed-origin blocks. * W7/W8: 0.3/100K inter-wave troughs exceed threshold (false early detection).

### Supplementary Methods: LLM Prompt Structure

The following prompt was used for the cumulative LLM condition (non-cumulative: HISTORICAL section omitted).

─────

You are an epidemiologist. A new COVID-19 wave ({wave}, {variant}) has just started in Japan. You only have data from the FIRST {N} DAYS (from K′-onset confirmation, 14 consecutive days of K′ > 0).

Japan has 11 blocks: Hokkaido, Tohoku, Kanto, Koshinetsu, Hokuriku, Tokai, Kinki, Chugoku, Shikoku, Kyushu, Okinawa.

**[CUMULATIVE ONLY] HISTORICAL CONFIRMED WAVE DATA:**

--- Wave k ({variant}): K′-onsets {regions, days}. Confirmed origins: {origins}.

**CURRENT WAVE first {N} days (per 100K):**

{region}: {rate}/100K, onset_day={day}, trend: rising/stable/falling Identify TOP 3 origin blocks. Respond ONLY with JSON:

{“top3_regions”:[“R1”,”R2”,”R3”],”reasoning”:”brief”}

─────

API: claude-haiku-4-5-20251001; max_tokens=300; temperature=1.0. Stateless; 3 independent runs per wave–timepoint–condition; mean F1 reported.

